# A scoping review protocol examining the application of large language models in healthcare education and public health learning spaces

**DOI:** 10.1101/2025.01.22.25320953

**Authors:** Henry Ndukwe, Emmanuel Otukpa

## Abstract

**Objective:** Through this scoping review, we aim to explore and synthesize existing knowledge and evidence on the learning approaches for incorporating LLMs into healthcare education and public health learning spaces. Specifically, we will attempt to investigate methods for auditing prompts for accuracy; tailoring prompts to improve task-specific accuracy and utility; and exploring how end-user feedback is used to refine and optimize LLM prompts over time. This review will provide a comprehensive understanding of how LLMs are being tailored and improved in these fields, contributing to the development of evidence-based strategies for their implementation. It will also identify areas for future research and innovation.

**Introduction:** The increasing integration of large language models (LLMs) into healthcare and public health practice and research highlights their potential to revolutionize service delivery, decision-making, and patient care. Despite these advancements, understanding how LLMs can be effectively tailored, audited, and refined for healthcare-specific tasks remains a critical area of inquiry. Key issues include, the accuracy of generated information, and their relevance to the medical and public health fields.

**Inclusion criteria:** Inclusion criteria will focus on studies addressing LLM applications in healthcare and public health, prompt engineering techniques, prompt auditing methods, and processes geared towards integrating user feedback. Articles that do not focus on healthcare or public health contexts and lack relevance to LLM learning approaches will be excluded.

**Methods:** The review is guided by the JBI methodology for scoping reviews complemented by updates from Levac et al. Databases including PubMed, Scopus, IEEE Xplore, and Web of Science will be searched for peer-reviewed articles, conference proceedings, and grey literature published in English and French from 2015 to 2025. Data extraction will include information on study characteristics, LLM models, prompt engineering strategies, auditing methodologies, and user feedback mechanisms. We will synthesize to identify trends, gaps, and best practices in leveraging LLMs to generate baseline data for auditing prompts that optimize AI learning and education needs in the healthcare and public health sector.

## Introduction

### Background and context

Large language models (LLMs) represent a novel innovation in computing and the part of what constitutes reactive artificial intelligence, offering the potential to revolutionize healthcare and public health through its natural language processing and language generation [1]. These models, exemplified by Generative Pre-trained Transformers (GPT) and its counterparts, can process and generate human-like text, enabling applications rooted in clinical decision-making, patient education, administrative efficiency, and medical and public health research [2,3]. As healthcare systems increasingly explore digital solutions to enhance care delivery, the role of LLMs is becoming more prominent. However, maximizing their potential while ensuring reliability and safety requires a nuanced understanding of their integration into healthcare-specific contexts [4].

The integration of LLMs into healthcare and public health education presents both opportunities and challenges. On the one hand, LLMs offer unprecedented capabilities for personalized learning, real-time clinical decision support, and enhanced access to medical knowledge and public health data [5]. They can assist in creating customized educational content, simulating clinical scenarios, and providing immediate feedback to learners using artificial intelligence (AI) models. And on the other hand, LLMs may help address the growing burden of healthcare-information overload and inaccurate online searching by synthesizing complex public and medical health prompts to illicit accurate and digestible output formats [4,5].

Despite these potential benefits, several challenges arise when considering the incorporation of LLMs into healthcare and public health education. The traditional learning framework has a shifting focus from scarcity towards the ubiquity of information via LLMs; fostering trust that would ensure user safety on a continuous basis. Future strategies that optimize efficiency with the leverage of AI-driven tools would require adaptable mechanism of feedback and continuous improvement. Ethical concerns surrounding data privacy, bias in AI systems, and the risk of over-reliance on technology must be carefully addressed [4,6]. There are also questions around the quality and accuracy of information generated by LLMs, particularly in rapidly evolving (as well as niche) medical fields [6]. Furthermore, educators and policymakers face the task of developing appropriate frameworks for integrating LLMs into curricula for learning and teaching or clinical practice modules [6,7].

### Challenges and gaps in knowledge

Despite the promise of having a standard output scoring system for generative AI prompts, challenges remain in deploying LLMs effectively in healthcare learning space [8]. Key concerns include the accuracy of generated outputs, biases embedded in models, and their contextual relevance to specific healthcare needs [8,9]. Addressing these challenges necessitates the development of precise prompt engineering techniques to tailor LLM responses for specialized tasks. Furthermore, auditing LLM outputs for accuracy, fairness, and effectiveness is critical to building trust and ensuring equitable healthcare delivery [10,11]. However, the methodologies and best practices for conducting such audits in healthcare and public health contexts are not well-documented [11,12].

### Role of user feedback

User feedback, particularly from healthcare and allied professionals, plays a vital role in refining LLM performance[14]. Feedback mechanisms provide insights into how LLMs can better align with clinical workflows and patient care priorities. While some studies have explored integrating user feedback in LLM applications, there remains a lack of clarity on how this process is operationalized and its impact on improving model outputs over time [10,14].

### Rationale for the Scoping review

Given the rapid evolution of LLMs and their potential adoption in healthcare, a comprehensive synthesis of existing knowledge is essential to understand ways for proper prompt engineering for generative AI. The ubiquitous nature of generative AI output poses a persistent risk of misinformation premised around LLMs’ learning algorithms, which are built to optimize outputs based on user input (prompts) [15,16]. It is crucial to understand possible oversight strategies to mitigate AI-information overload in learning environments. To that end, this review will explore how LLMs have been deployed and audited in healthcare education and public health learning spaces. By investigating the methods used to audit LLM prompts; strategies to enhance task-specific accuracy and utility, and the integration of user feedback can be proposed to refine LLM outputs within healthcare, clinical and public health settings.

### Objectives

In this scoping review, we aim to explore the current state of LLM integration in healthcare and public health education, identify best practices, and highlight areas requiring further research and development. We seek to provide actionable insights for researchers, healthcare professionals, and policymakers to optimize the use of LLMs in healthcare and public health education. The identification of best practices and knowledge gaps, it will contribute to advancing the safe and effective implementation of LLM technologies in these critical fields

### Review questions

1. What methods are used to audit prompts for accuracy in healthcare and public health LLMs?

2. How are prompts tailored to specific tasks to improve the accuracy and utility of LLM outputs?

3. How have end-user feedback been integrated into the process of refining LLM prompts over time?

## Materials and Methods

We intend to conduct this scoping review in accordance with the JBI methodology for scoping reviews [17]. The methodology entails a systematic approach to searching, screening, and reporting that include the following stages: (1) identification of the research question (s); (2) identification of relevant databases and studies; (3) selection of studies; (4) data extraction; (5) interpretation, summarization and dissemination of the results.

### Inclusion criteria

We intend to focus on studies whose focus is on the application or development of large language models (LLMs) usage and application/integration in healthcare or public health contexts. Examples of relevant contexts include clinical decision support, patient education, administrative processes, and public health interventions and education modules. In terms of the scope of LLM integration, the studies must explore learning approaches for integrating LLMs, including prompt engineering, auditing methodologies, and user feedback mechanisms.

Specific publication inclusion criteria will focus on peer-reviewed articles, conference proceedings, and grey literature (e.g., reports, white papers). Additionally, all study designs are eligible, including experimental, observational, qualitative, and mixed-methods studies. Studies focusing on frameworks, methodologies, and case studies will also be considered. We will also consider any study published in English or French language. And with a timeframe for inclusion from 2015 to 2025, reflecting a period of rapid advancements in LLM technology and its applications.

In terms of relevance, studies must address at least one of the following core areas:

- Methods for auditing LLM prompt for accuracy, fairness, and effectiveness.
- Techniques for tailoring LLM prompts to healthcare-specific tasks.
- Approaches for integrating user feedback from healthcare professionals to refine and improve LLM outputs.

### Search strategy

Our search strategy will aim to locate both published and unpublished studies. A three-step search strategy will be utilized in this review. First, we will conduct an initial limited search of MEDLINE (PubMed) and CINAHL (EBSCO) to identify articles on the topic. The text words contained in the titles and abstracts of relevant articles, and the index terms used to describe the articles will be used to develop a full search strategy for reporting the name of the relevant databases/information sources *(see Appendix 1*). The search strategy, including all identified keywords and index terms, will be adapted for each included database and/or information source. The reference list of all included sources of evidence will be screened for additional studies. We will search relevant peer-reviewed, English and French-language articles published between January 1, 2015, and January 31, 2025, without methodological restrictions, in several electronic databases, as well as sources with broad specificity (Web of Science and Google Scholar). Searches will extend to grey literature sources, including institutional reports, white papers, and preprints on platforms like arXiv.

### Source of evidence selection (databases)

Searches will be conducted in electronic databases for bibliographic sources including Medline, Embase, and Scopus. We will include varied electronic data sources including Web of Science, Google Scholar and IEEE Xplore.

### Search terms

We will employ adjacency search and combination of keywords and Medical Subject Headings (MeSH) terms will be used, including: *“large language models”, “prompt engineering”, “healthcare”, “public health”, “auditing methods”, “user feedback”, “artificial intelligence”*. The Boolean operators (AND, OR, NOT) will combine the above terms to refine quantity and quality of search hits. A detailed search strategy is provided in Appendix 1.

### Study selection

We will use the online tool Covidence®, which allows for simultaneous title, abstract and full text article reviews; to screen through articles to export included titles to Excel ® for analysis. Two researchers will independently assess articles for inclusion by screening the titles, abstracts, and full texts of studies returned through the search process. Where there are disagreements between the two independent reviewers on the eligibility of a paper for inclusion, a third reviewer will adjudicate using same inclusion criteria to resolve the conflict.

### Data extraction

We will use a standardized data charting form *(Appendix 2)* to extract relevant data from included studies. The following details will be extracted:

1. **Study Characteristics**: Title, authors, year, and country of publication.
2. **LLM Details**: Type of LLM, application context, and specific tasks addressed.
3. **Methodologies**: Techniques for prompt engineering, auditing methods, and feedback mechanisms.
4. **Outcomes**: Measures of effectiveness, fairness, and task-specific utility.
5. **User Feedback**: Processes for incorporating feedback from healthcare professionals and the impact on model refinement.

### Data synthesis and presentation

We will analyze the data using descriptive statistics and thematic analysis, with results organized in tables and charts and presented into themes that reflect the review objectives. Our thematic analysis will identify patterns and trends in the application of LLMs.

Quantitative data, where applicable, will be summarized descriptively. We will also conduct a narrative synthesis to integrate findings across studies, focusing on methods, outcomes, and identified gaps.

### Ethics and dissemination

Ethical approval is not required because primary data collection is not involved in this study but rather analyzing both published and grey literature. However, the findings of this study will be disseminated through peer-reviewed publications and conferences as well as in relevant stakeholder fora. In case of any amendments to the protocol following its publication, we will provide the date of each amendment, describe the change(s), and report the rationale for the change(s) in future publications arising from this protocol.

### Strengths and limitations

The strength of this review lies within the systematic approach to synthesizing the diverse evidence on the integration of large language models (LLMs) in healthcare, clinical and public health, with a focus on prompt engineering, auditing, and user feedback mechanisms, which, is a relatively niche concept. By utilizing a broad range of sources, including peer-reviewed studies and grey literature, the review will provide a comprehensive understanding of current practices, trends, and gaps in the field. Its focus on healthcare-specific applications ensures relevance to real-world policy relevant challenges, while the inclusion of feedback mechanisms highlights its alignment with user-centered design principles. Conversely, potential limitations include the restriction to English and French language publications, which may exclude relevant studies in other languages, and reliance on available literature that may underrepresent unpublished or proprietary methods used by private collectives. Additionally, the rapidly evolving nature of LLM technologies means that findings may quickly become outdated, necessitating continuous updates to maintain relevance.

## Data Availability

N/A

## Acknowledgements

The authors would like to acknowledge Griffith University, and the African Population and Health Research Center(APHRC) for their institutional support towards this innovative collaboration.

## Declarations

None to declare

## Author contributions

HN and EO initiated the conception of the review and EO led the drafting of the protocol while HN provided critical review.

## Appendices

### Appendix 1 Search strategy

#### Database Search Strategy

We will use the advance search builder of the PubMed and CINAHL databases, which allows for the use of wildcards (*) and lengthy search terms. We limited our search to a period between 1^st^ January 2015 to 31^st^ January 2025.

**Table 1.** PubMed Search Strategy.

|  |  |
| --- | --- |
| #1 | "Large Language Models" OR LLMs OR GPT-3 OR GPT-4 OR BERT OR RoBERTa OR T5 OR MedPaLM OR BioGPT OR ClinicalBERT OR pubmedBERT OR BioMegatron OR GatorTron OR ALBERT OR CODEX OR Glalactica OR SapBERT OR MIMIC-BERT |
| #2 | "Prompt Engineering" OR "Natural language processing" OR "machine learning" OR "deep learning" OR "language models" OR prompts OR fine-tuning |
| #3 | Medicine OR disease OR epidemiology OR "public health" OR "healthcare systems" OR "healthcare delivery"OR"auditing methods"OR "user feedback", |
| #4 | **Diagnosis OR treatment OR "drug discovery" OR "medical imaging" OR health OR Application OR education OR "disease surveillance"** |
| #5 | #1 AND #4 |
| #6 | #1 AND #2 AND #4 |
| #7 | #1 AND #2 AND #3 AND #4 |

### Appendix 2 Data extraction instrument

**Appendix 2: Data extraction instrument**
| Key domain | Data point | Description |
| --- | --- | --- |
| Study Identification | Title | Indicates the study title |
|  | Authors | Indicates the names of the authors |
|  | Publication year |  |
|  | Country /region | Indicates the country of study and geographical region |
|  | Source | This indicates Database, journal, or organization where the study was found |
| Study characteristics | Study type | Type of study (e.g., experimental, observational, qualitative, case study) |
|  | Objective | Primary aim or research question of the study. |
|  | Context | Description of the healthcare or public health setting |
| LLM detail | LLM type | Name and version of the large language model used (e.g., GPT, BERT) |
|  | Application | Specific application of the LLM in healthcare or public health (e.g., clinical decision-making). |
|  | Task specific use | Specific tasks for which the LLM was used or tailored. |
| Prompt Engineering details | Techniques | Methods used to design and refine prompts for specific tasks. |
|  | Tailoring/Customizing strategies | Techniques for tailoring prompts to improve accuracy and utility. |
|  | Auditing process | Methods used to assess LLM outputs for accuracy, fairness, and effectiveness. |
|  | Auditing outcomes | Key findings or metrics from the auditing process. |
| End-user feedback | Feedback processes and mechanism | Processes for gathering feedback from healthcare professionals. |
|  | Feedback implementation | Description of how feedback was used to refine prompts or LLM outputs. |
|  | Feedback impact assessment | Outcomes resulting from feedback integration. |
| Results and outcomes | Key findings | Major findings relevant to the study objectives |
|  | Effectiveness | Measures of success or effectiveness of LLM application |
|  | Challenges | Any reported challenges or limitations in the study. |
| Gaps and relevance | Relevance to review objectives | Description of how the study aligns with the review objectives. |
|  | Identified gaps | Gaps or unanswered questions highlighted by the study. |
| Quality assessment | Randomized Trials/Interventions | Cochrane Risk of Bias assessment tool |
|  | Observational studies | Newcastle-Ottawa scale (NOS) for non-randomized studies |
|  | Qualitative studies | JBICritical Appraisal Checklist |
|  | Study limitations | Limitations or biases reported by the study authors |
|  | Reviewer observations | Any additional domains or observations identified by the reviewer. |

